# Comparison of influenza and COVID-19–associated hospitalizations among children < 18 years old in the United States — FluSurv-NET (October–April 2017–2021) and COVID-NET (October 2020–September 2021)

**DOI:** 10.1101/2022.03.09.22271788

**Authors:** Miranda J. Delahoy, Dawud Ujamaa, Christopher A. Taylor, Charisse Cummings, Onika Anglin, Rachel Holstein, Jennifer Milucky, Alissa O’Halloran, Kadam Patel, Huong Pham, Michael Whitaker, Arthur Reingold, Shua J. Chai, Nisha B. Alden, Breanna Kawasaki, James Meek, Kimberly Yousey-Hindes, Evan J. Anderson, Kyle P. Openo, Andy Weigel, Kenzie Teno, Libby Reeg, Lauren Leegwater, Ruth Lynfield, Melissa McMahon, Susan Ropp, Dominic Rudin, Alison Muse, Nancy Spina, Nancy M. Bennett, Kevin Popham, Laurie M. Billing, Eli Shiltz, Melissa Sutton, Ann Thomas, William Schaffner, H. Keipp Talbot, Melanie T. Crossland, Keegan McCaffrey, Aron J. Hall, Erin Burns, Meredith McMorrow, Carrie Reed, Fiona P. Havers, Shikha Garg

## Abstract

**Background:** Influenza virus and SARS-CoV-2 are significant causes of respiratory illness in children.

**Methods:** Influenza and COVID-19-associated hospitalizations among children <18 years old were analyzed from FluSurv-NET and COVID-NET, two population-based surveillance systems with similar catchment areas and methodology. The annual COVID-19-associated hospitalization rate per 100 000 during the ongoing COVID-19 pandemic (October 1, 2020–September 30, 2021) was compared to influenza-associated hospitalization rates during the 2017–18 through 2019–20 influenza seasons. In-hospital outcomes, including intensive care unit (ICU) admission and death, were compared.

**Results:** Among children <18 years old, the COVID-19-associated hospitalization rate (48.2) was higher than influenza-associated hospitalization rates: 2017–18 (33.5), 2018–19 (33.8), and 2019–20 (41.7). The COVID-19-associated hospitalization rate was higher among adolescents 12–17 years old (COVID-19: 59.9; influenza range: 12.2-14.1), but similar or lower among children 5–11 (COVID-19: 25.0; influenza range: 24.3-31.7) and 0–4 (COVID-19: 66.8; influenza range: 70.9-91.5) years old. Among children <18 years old, a higher proportion with COVID-19 required ICU admission compared with influenza (26.4% vs 21.6%; p<0.01). Pediatric deaths were uncommon during both COVID-19- and influenza-associated hospitalizations (0.7% vs 0.5%; p=0.28).

**Conclusions:** In the setting of extensive mitigation measures during the COVID-19 pandemic, the annual COVID-19-associated hospitalization rate during 2020–2021 was higher among adolescents and similar or lower among children <12 years old compared with influenza during the three seasons before the COVID-19 pandemic. COVID-19 adds substantially to the existing burden of pediatric hospitalizations and severe outcomes caused by influenza and other respiratory viruses.

**Summary:** Annual hospitalization rates and proportions of hospitalized children experiencing severe outcomes were as high or higher for COVID-19 during October 2020–September 2021 compared with influenza during the three seasons before the COVID-19 pandemic, based on U.S. population-based surveillance data.

## Introduction

Influenza virus and SARS-CoV-2 are significant causes of respiratory illness and can lead to severe illness including death in children [1–5]. Annual influenza vaccination is approved and recommended for all individuals ≥ 6 months old without contraindications [6], whereas COVID-19 vaccines are currently authorized or approved for persons ≥ 5 years old [7]. Influenza-associated hospitalization rates are typically highest among adults aged ≥ 65 years, followed during some seasons by adults aged 50–64 years and during other seasons by children 0–4 years old [8]. COVID-19-associated hospitalization rates are similarly higher among adults compared with children [9]. However, data comparing influenza versus COVID-19-associated hospitalizations among children are limited [10,11]. Such data are useful for evaluating the impact of mitigation measures and for interpreting disease burden measures, which can provide useful context to inform COVID-19 vaccine recommendations for children < 5 years old. We compared hospitalization rates, clinical characteristics, and outcomes among children < 18 years old hospitalized with seasonal influenza or COVID-19 in the United States.

## Methods

The Influenza Hospitalization Surveillance Network (FluSurv-NET) [8] and the Coronavirus Disease 2019-Associated Hospitalization Surveillance Network (COVID-NET) [12,13] conduct population-based surveillance for laboratory-confirmed influenza-associated and COVID-19-associated hospitalizations, respectively. FluSurv-NET surveillance among children was initiated during the 2003–04 influenza season. COVID-NET surveillance was initiated in March 2020 using FluSurv-NET infrastructure [12]. During the 2017–18 through 2020–21 influenza seasons, FluSurv-NET surveillance was conducted in counties in 14 states participating in either the Emerging Infections Program (California, Colorado, Connecticut, Georgia, Maryland [Baltimore Metropolitan Area], Minnesota, New Mexico, New York, Oregon, and Tennessee) or the Influenza Hospitalization Surveillance Project (Iowa [2020–21 season only], Michigan, Ohio, and Utah), with a total catchment population of approximately 29 million persons (9% of the U.S. population). COVID-NET conducts surveillance in all FluSurv-NET counties and all counties in Maryland, with a catchment population of approximately 32 million persons (10% of the U.S. population).

As previously described [8,13], FluSurv-NET surveillance is conducted during each influenza season (October 1 through April 30) and COVID-NET surveillance is conducted year-round. A FluSurv-NET or COVID-NET case is defined as a hospitalized patient who is a resident of the FluSurv-NET or COVID-NET catchment area, with a positive influenza test (rapid antigen detection, molecular assay, direct or indirect immunofluorescence assay, or viral culture) or SARS-CoV-2 test (rapid antigen detection or molecular assay) during or ≤ 14 days before hospitalization. Testing for influenza virus or SARS-CoV-2 is performed at the discretion of health care practitioners or according to hospital testing practices. Trained surveillance staff identify all catchment area residents hospitalized with laboratory-confirmed influenza or COVID-19 using laboratory, hospital, and reportable conditions databases.

Medical record abstractions are conducted using standardized data collection forms to obtain information on demographics, clinical characteristics, interventions (invasive mechanical ventilation and extracorporeal membrane oxygenation), and outcomes (intensive care unit [ICU] admission, pneumonia, and in-hospital death from any cause) during an influenza- or COVID-19-associated hospitalization.

Obesity status was determined using body mass index (≥ 95th percentile for sex and age), ICD-10-CM discharge diagnosis codes, and problem lists from medical records for non-pregnant persons ≥ 2 years old. Acute symptoms at admission were abstracted from history and physical exam notes; the list of abstracted symptoms varied by surveillance platform and year. Acute respiratory or febrile symptoms (fever, congestion/runny nose, cough, shortness of breath, sore throat, upper respiratory illness or influenza-like illness, and wheezing) were abstracted for all FluSurv-NET seasons and for COVID-NET. Additional symptoms abstracted for FluSurv-NET by season and for COVID-NET are detailed in Table 1 and Supplementary Table 1.

**Table 1.**
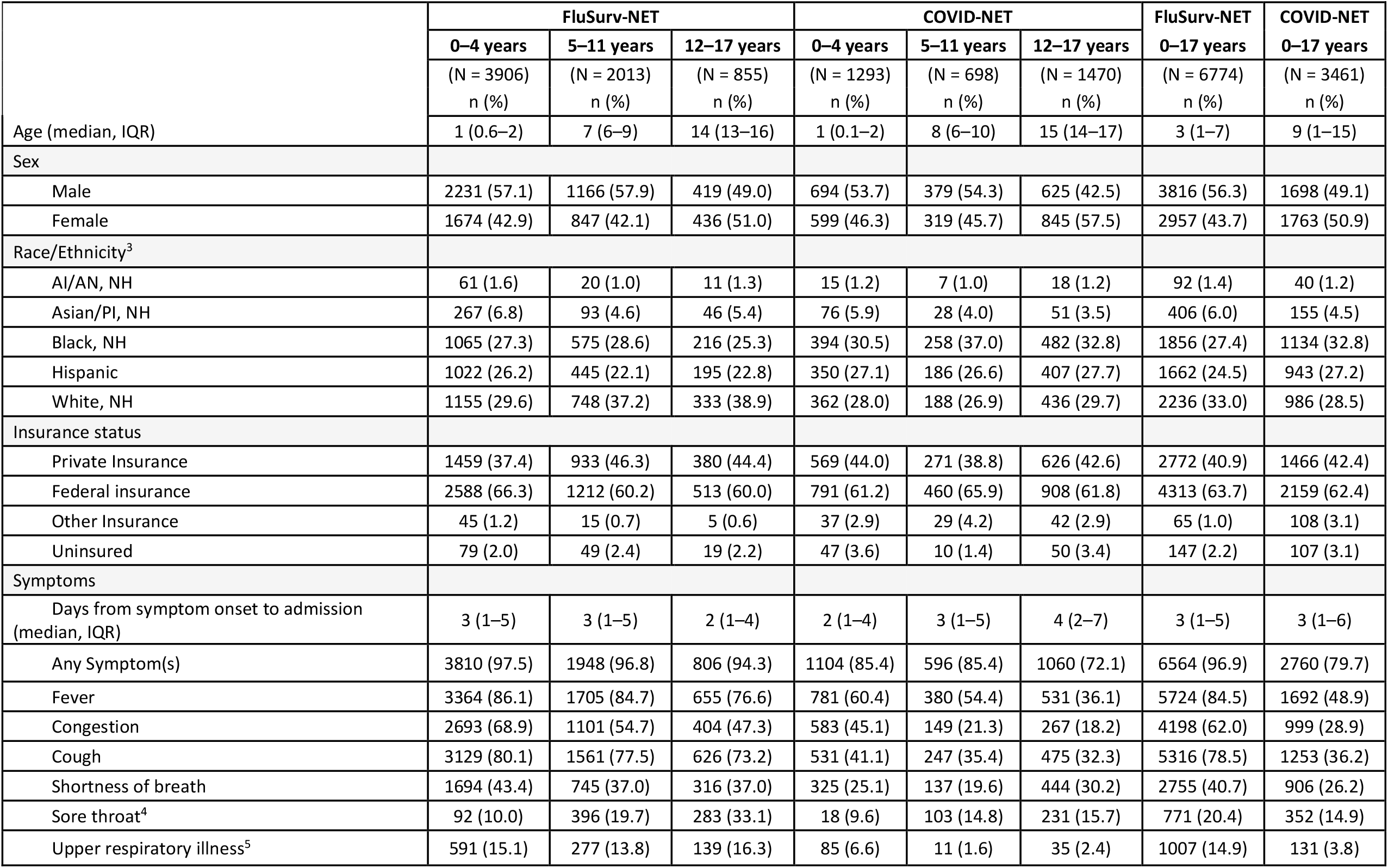
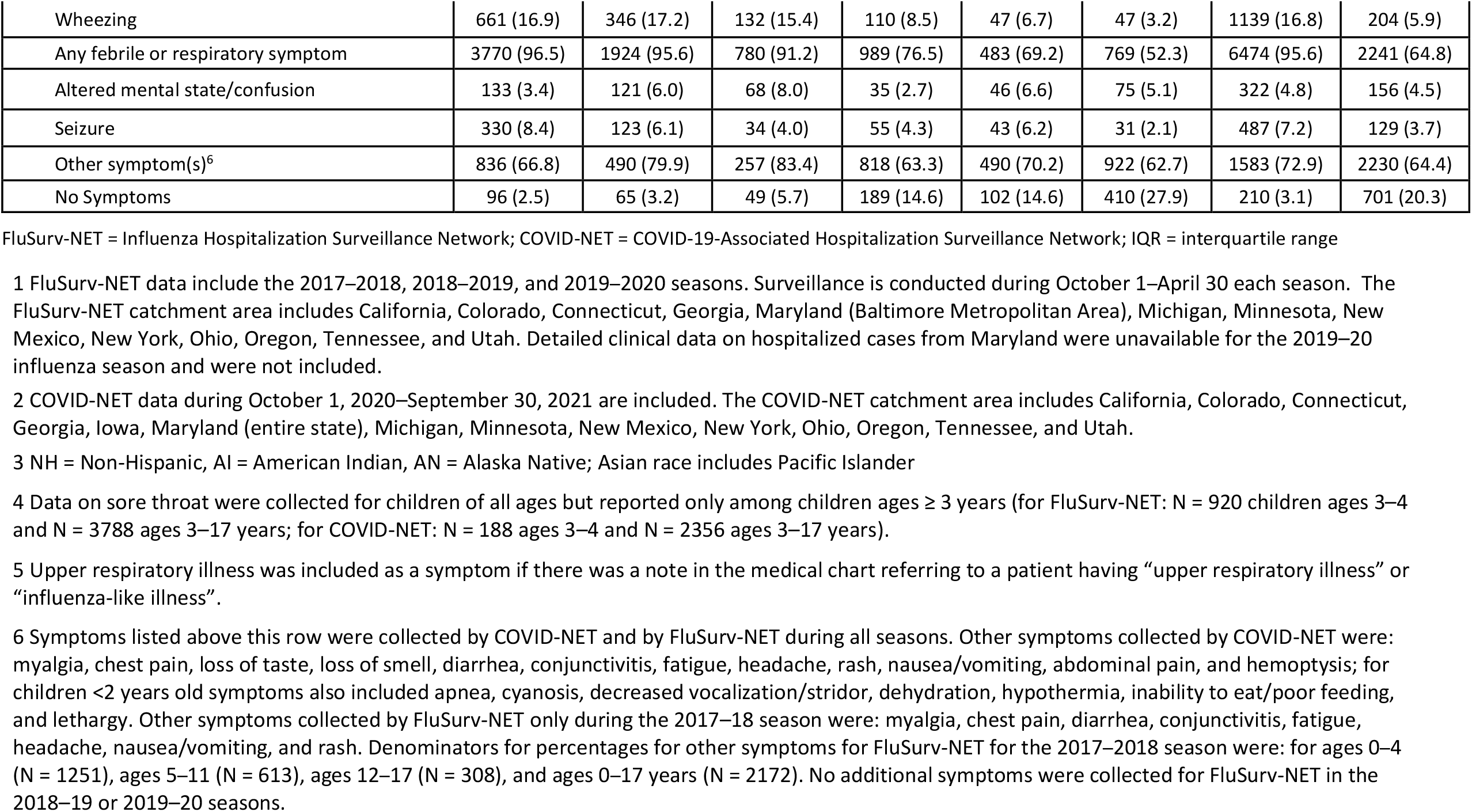
Demographic and clinical characteristics of children <18 years old hospitalized with influenza or COVID-19 – FluSurv-NET^1^ and COVID-NET^2^.

|  | FluSurv-NET |  |  | COVID-NET |  |  | FluSurv-NET | COVID-NET |
| --- | --- | --- | --- | --- | --- | --- | --- | --- |
|  | 0–4 years | 5–11 years | 12–17 years | 0–4 years | 5–11 years | 12–17 years | 0–17 years | 0–17 years |
|  | (N = 3906)<br>n (%) | (N = 2013)<br>n (%) | (N = 855)<br>n (%) | (N = 1293)<br>n (%) | (N = 698)<br>n (%) | (N = 1470)<br>n (%) | (N = 6774)<br>n (%) | (N = 3461)<br>n (%) |
| Age (median, IQR) | 1 (0.6–2) | 7 (6–9) | 14 (13–16) | 1 (0.1–2) | 8 (6–10) | 15 (14–17) | 3 (1–7) | 9 (1–15) |
| Sex |  |  |  |  |  |  |  |  |
| Male | 2231 (57.1) | 1166 (57.9) | 419 (49.0) | 694 (53.7) | 379 (54.3) | 625 (42.5) | 3816 (56.3) | 1698 (49.1) |
| Female | 1674 (42.9) | 847 (42.1) | 436 (51.0) | 599 (46.3) | 319 (45.7) | 845 (57.5) | 2957 (43.7) | 1763 (50.9) |
| Race/Ethnicity<sup>3</sup> |  |  |  |  |  |  |  |  |
| AI/AN, NH | 61 (1.6) | 20 (1.0) | 11 (1.3) | 15 (1.2) | 7 (1.0) | 18 (1.2) | 92 (1.4) | 40 (1.2) |
| Asian/PI, NH | 267 (6.8) | 93 (4.6) | 46 (5.4) | 76 (5.9) | 28 (4.0) | 51 (3.5) | 406 (6.0) | 155 (4.5) |
| Black, NH | 1065 (27.3) | 575 (28.6) | 216 (25.3) | 394 (30.5) | 258 (37.0) | 482 (32.8) | 1856 (27.4) | 1134 (32.8) |
| Hispanic | 1022 (26.2) | 445 (22.1) | 195 (22.8) | 350 (27.1) | 186 (26.6) | 407 (27.7) | 1662 (24.5) | 943 (27.2) |
| White, NH | 1155 (29.6) | 748 (37.2) | 333 (38.9) | 362 (28.0) | 188 (26.9) | 436 (29.7) | 2236 (33.0) | 986 (28.5) |
| Insurance status |  |  |  |  |  |  |  |  |
| Private Insurance | 1459 (37.4) | 933 (46.3) | 380 (44.4) | 569 (44.0) | 271 (38.8) | 626 (42.6) | 2772 (40.9) | 1466 (42.4) |
| Federal insurance | 2588 (66.3) | 1212 (60.2) | 513 (60.0) | 791 (61.2) | 460 (65.9) | 908 (61.8) | 4313 (63.7) | 2159 (62.4) |
| Other Insurance | 45 (1.2) | 15 (0.7) | 5 (0.6) | 37 (2.9) | 29 (4.2) | 42 (2.9) | 65 (1.0) | 108 (3.1) |
| Uninsured | 79 (2.0) | 49 (2.4) | 19 (2.2) | 47 (3.6) | 10 (1.4) | 50 (3.4) | 147 (2.2) | 107 (3.1) |
| Symptoms |  |  |  |  |  |  |  |  |
| Days from symptom onset to admission (median, IQR) | 3 (1–5) | 3 (1–5) | 2 (1–4) | 2 (1–4) | 3 (1–5) | 4 (2–7) | 3 (1–5) | 3 (1–6) |
| Any Symptom(s) | 3810 (97.5) | 1948 (96.8) | 806 (94.3) | 1104 (85.4) | 596 (85.4) | 1060 (72.1) | 6564 (96.9) | 2760 (79.7) |
| Fever | 3364 (86.1) | 1705 (84.7) | 655 (76.6) | 781 (60.4) | 380 (54.4) | 531 (36.1) | 5724 (84.5) | 1692 (48.9) |
| Congestion | 2693 (68.9) | 1101 (54.7) | 404 (47.3) | 583 (45.1) | 149 (21.3) | 267 (18.2) | 4198 (62.0) | 999 (28.9) |
| Cough | 3129 (80.1) | 1561 (77.5) | 626 (73.2) | 531 (41.1) | 247 (35.4) | 475 (32.3) | 5316 (78.5) | 1253 (36.2) |
| Shortness of breath | 1694 (43.4) | 745 (37.0) | 316 (37.0) | 325 (25.1) | 137 (19.6) | 444 (30.2) | 2755 (40.7) | 906 (26.2) |
| Sore throat<sup>4</sup> | 92 (10.0) | 396 (19.7) | 283 (33.1) | 18 (9.6) | 103 (14.8) | 231 (15.7) | 771 (20.4) | 352 (14.9) |
| Upper respiratory illness<sup>5</sup> | 591 (15.1) | 277 (13.8) | 139 (16.3) | 85 (6.6) | 11 (1.6) | 35 (2.4) | 1007 (14.9) | 131 (3.8) |
| Wheezing | 661 (16.9) | 346 (17.2) | 132 (15.4) | 110 (8.5) | 47 (6.7) | 47 (3.2) | 1139 (16.8) | 204 (5.9) |
| Any febrile or respiratory symptom | 3770 (96.5) | 1924 (95.6) | 780 (91.2) | 989 (76.5) | 483 (69.2) | 769 (52.3) | 6474 (95.6) | 2241 (64.8) |
| Altered mental state/confusion | 133 (3.4) | 121 (6.0) | 68 (8.0) | 35 (2.7) | 46 (6.6) | 75 (5.1) | 322 (4.8) | 156 (4.5) |
| Seizure | 330 (8.4) | 123 (6.1) | 34 (4.0) | 55 (4.3) | 43 (6.2) | 31 (2.1) | 487 (7.2) | 129 (3.7) |
| Other symptom(s)<sup>6</sup> | 836 (66.8) | 490 (79.9) | 257 (83.4) | 818 (63.3) | 490 (70.2) | 922 (62.7) | 1583 (72.9) | 2230 (64.4) |
| No Symptoms | 96 (2.5) | 65 (3.2) | 49 (5.7) | 189 (14.6) | 102 (14.6) | 410 (27.9) | 210 (3.1) | 701 (20.3) |

FluSurv-NET = Influenza Hospitalization Surveillance Network; COVID-NET = COVID-19-Associated Hospitalization Surveillance Network; IQR = interquartile range
1 FluSurv-NET data include the 2017–2018, 2018–2019, and 2019–2020 seasons. Surveillance is conducted during October 1–April 30 each season. The FluSurv-NET catchment area includes California, Colorado, Connecticut, Georgia, Maryland (Baltimore Metropolitan Area), Michigan, Minnesota, New Mexico, New York, Ohio, Oregon, Tennessee, and Utah. Detailed clinical data on hospitalized cases from Maryland were unavailable for the 2019–20 influenza season and were not included.
2 COVID-NET data during October 1, 2020–September 30, 2021 are included. The COVID-NET catchment area includes California, Colorado, Connecticut, Georgia, Iowa, Maryland (entire state), Michigan, Minnesota, New Mexico, New York, Ohio, Oregon, Tennessee, and Utah.
NH = Non-Hispanic, AI = American Indian, AN = Alaska Native; Asian race includes Pacific Islander
4 Data on sore throat were collected for children of all ages but reported only among children ages ≥ 3 years (for FluSurv-NET: N = 920 children ages 3–4 and N = 3788 ages 3–17 years; for COVID-NET: N = 188 ages 3–4 and N = 2356 ages 3–17 years).
5 Upper respiratory illness was included as a symptom if there was a note in the medical chart referring to a patient having “upper respiratory illness” or “influenza-like illness”.
6 Symptoms listed above this row were collected by COVID-NET and by FluSurv-NET during all seasons. Other symptoms collected by COVID-NET were: myalgia, chest pain, loss of taste, loss of smell, diarrhea, conjunctivitis, fatigue, headache, rash, nausea/vomiting, abdominal pain, and hemoptysis; for children <2 years old symptoms also included apnea, cyanosis, decreased vocalization/stridor, dehydration, hypothermia, inability to eat/poor feeding, and lethargy. Other symptoms collected by FluSurv-NET only during the 2017–18 season were: myalgia, chest pain, diarrhea, conjunctivitis, fatigue, headache, nausea/vomiting, and rash. Denominators for percentages for other symptoms for FluSurv-NET for the 2017–2018 season were: for ages 0–4 (N = 1251), ages 5–11 (N = 613), ages 12–17 (N = 308), and ages 0–17 years (N = 2172). No additional symptoms were collected for FluSurv-NET in the 2018–19 or 2019–20 seasons.

Monthly hospitalization counts were determined for influenza (October–April during the 2017–18 through 2020–21 influenza seasons) and COVID-19 (March 2020–September 2021). Unadjusted influenza and COVID-19-associated hospitalization rates per 100 000 children and 95% confidence intervals (CIs) were calculated by dividing the total number of hospitalizations by population denominators obtained from the National Center for Health Statistics postcensal bridged-race population estimates [14]. The COVID-19-associated hospitalization rate was calculated for a 1-year period (October 1, 2020–September 30, 2021). This annual rate was compared with influenza-associated hospitalization rates during October 1–April 30 of each of the 2017–18 through 2019–20 influenza seasons. Influenza occurs seasonally in the United States with low detection during May–September [15,16], suggesting few influenza-associated hospitalizations are missed outside the October–April surveillance window. Thus, influenza-associated hospitalization rates during October–April were used to represent annual rates. Weekly hospitalization rates per 100 000 children were calculated for influenza (overall and by influenza virus type) and COVID-19.

The frequencies of select characteristics and outcomes were calculated for children hospitalized with influenza (2017–18 through 2019-20 influenza seasons), and COVID-19 (October 1, 2020–September 30, 2021). *P* values were calculated using Pearson chi-square or Wilcoxon rank sum tests. Statistical significance was set at α = .05; all tests were 2-sided. Statistical analyses were performed in SAS version 9.4 (SAS Institute).

Children with laboratory-confirmed influenza or COVID-19 may have been hospitalized primarily for other reasons but found incidentally to have influenza or COVID-19. We conducted sensitivity analyses to determine whether severe outcomes occurring during hospitalizations differed by symptom status (presence of ≥ 1 symptom at admission) for both influenza and COVID-19 and by reason for admission for COVID-19 (these data were not available in FluSurv-NET). For COVID-NET, the primary reason for admission was determined based on the chief complaint and history of present illness from the hospital admission note (Supplementary Table 2).

FluSurv-NET and COVID-NET surveillance activities were reviewed by CDC and were conducted consistent with applicable federal law and CDC policy (e.g., 45 CFR. Part 46.102(l)(2), 21 CFR part 56; 42 USC. §241(d); 5 USC §552a; 44 USC §3501 et seq). Sites participating in FluSurv-NET and COVID-NET obtained human subjects and ethics approvals from their respective state and local health department and academic partner Institutional Review Boards as needed.

## Results

From October 2017 until February 2020, monthly counts of influenza-associated hospitalizations followed a typical seasonal pattern (Figure 1); during March–April 2020, monthly influenza counts decreased abruptly, coinciding with the release on March 16, 2020 of national guidance for slowing the spread of COVID-19, which included school closures and other mitigation measures [17]. Subsequently, during October 1, 2020–April 30, 2021, only 9 influenza-associated hospitalizations among children were reported to FluSurv-NET. Starting in March 2020, when COVID-NET surveillance was initiated, COVID-19-associated hospitalizations were identified in children each month (range: 22–470 hospitalizations per month during March 2020–September 2021).

**Figure 1.**
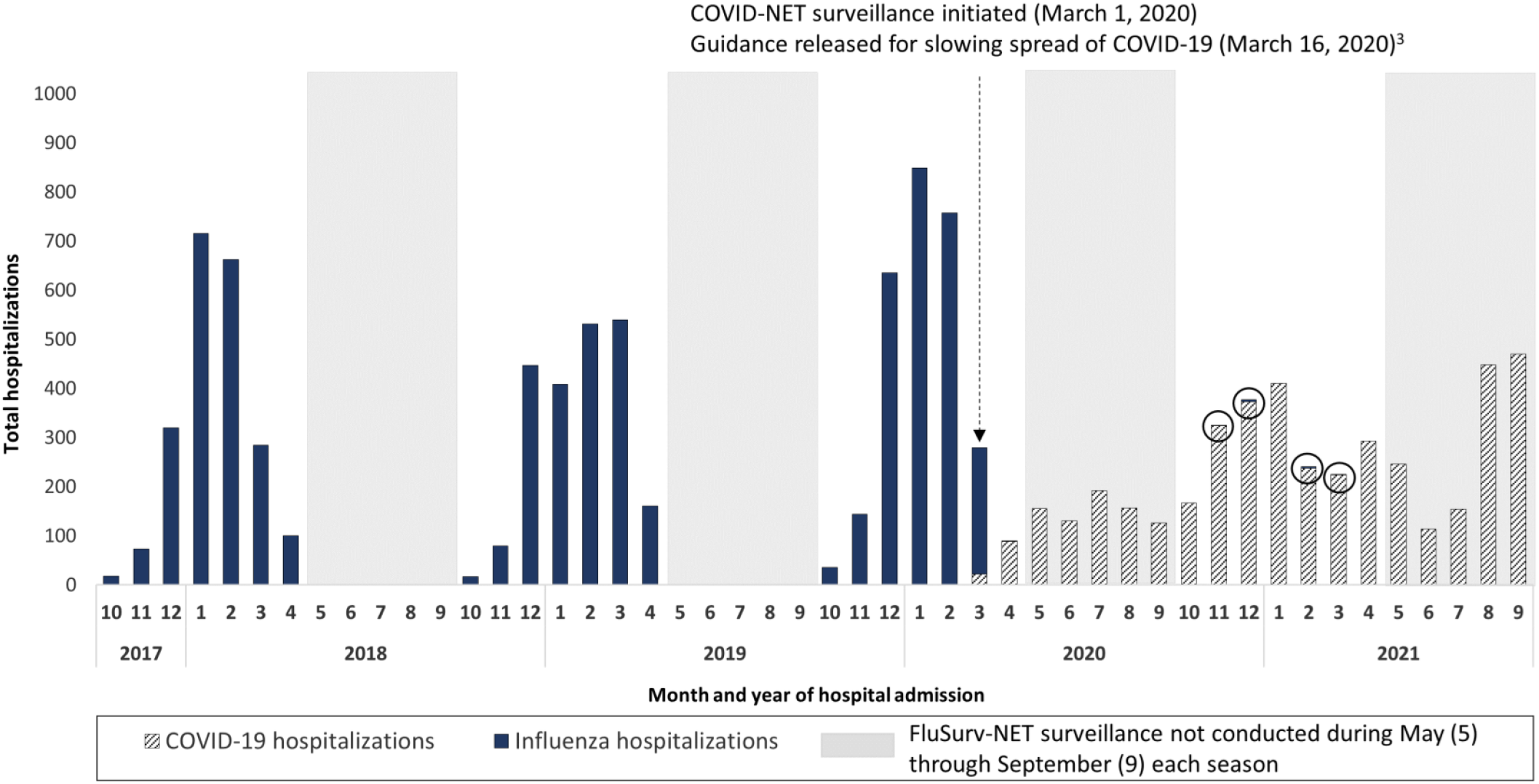
Counts of COVID-19- and influenza-associated hospitalizations by month among children <18 years old, FluSurv-NET^1^ and COVID-NET^2^, October 2017–September 2021. FluSurv-NET = Influenza Hospitalization Surveillance Network; COVID-NET = COVID-19-Associated Hospitalization Surveillance Network 1 The 9 influenza-associated hospitalizations reported to FluSurv-NET during October 1, 2020–April 30, 2021 are circled to improve visibility. The FluSurv-NET catchment area includes California, Colorado, Connecticut, Georgia, Iowa (2020–21 season only), Maryland (Baltimore Metropolitan Area), Michigan, Minnesota, New Mexico, New York, Ohio, Oregon, Tennessee, and Utah. 2 The COVID-NET catchment area includes California, Colorado, Connecticut, Georgia, Iowa, Maryland (entire state), Michigan, Minnesota, New Mexico, New York, Ohio, Oregon, Tennessee, and Utah. 3 Guidance for slowing the spread of COVID-19 was released on March 16, 2020, after which school closures began. See, e.g., https://www.cdc.gov/mmwr/volumes/69/wr/mm6915e2.htm.

Among all children, influenza-associated hospitalization rates observed during the three seasons prior to the COVID-19 pandemic (2017–18 through 2019–20; rate range = 33.5–41.7 per 100 000; 95% CI of highest-burden season [2019–20]: 40.2–43.3) were lower than one annual COVID-19-associated hospitalization rate observed during October 2020–September 2021 of the ongoing COVID-19 pandemic (48.2; 95% CI: 46.6–49.8) (Supplementary Table 3; Figure 2). However, differences were observed by age. Among children 0–4 years old, influenza-associated hospitalization rates for the 2017–18 season (71.0) and 2018-19 season (70.9) were similar to the annual COVID-19-associated hospitalization rate (66.8), but the influenza-associated hospitalization rate for the 2019-20 season (91.5) was higher. Influenza- and COVID-19-associated hospitalization rates were similar among children 5–11 years old (2017–18 through 2019–20 influenza seasons: 24.3–31.7; COVID-19: 25.0). Among adolescents (12–17 years old), influenza-associated hospitalization rates for the 2017–18 through 2019–20 seasons (rate range: 12.2–14.1) were lower than the COVID-19-associated hospitalization rate (59.9).

**Figure 2.**
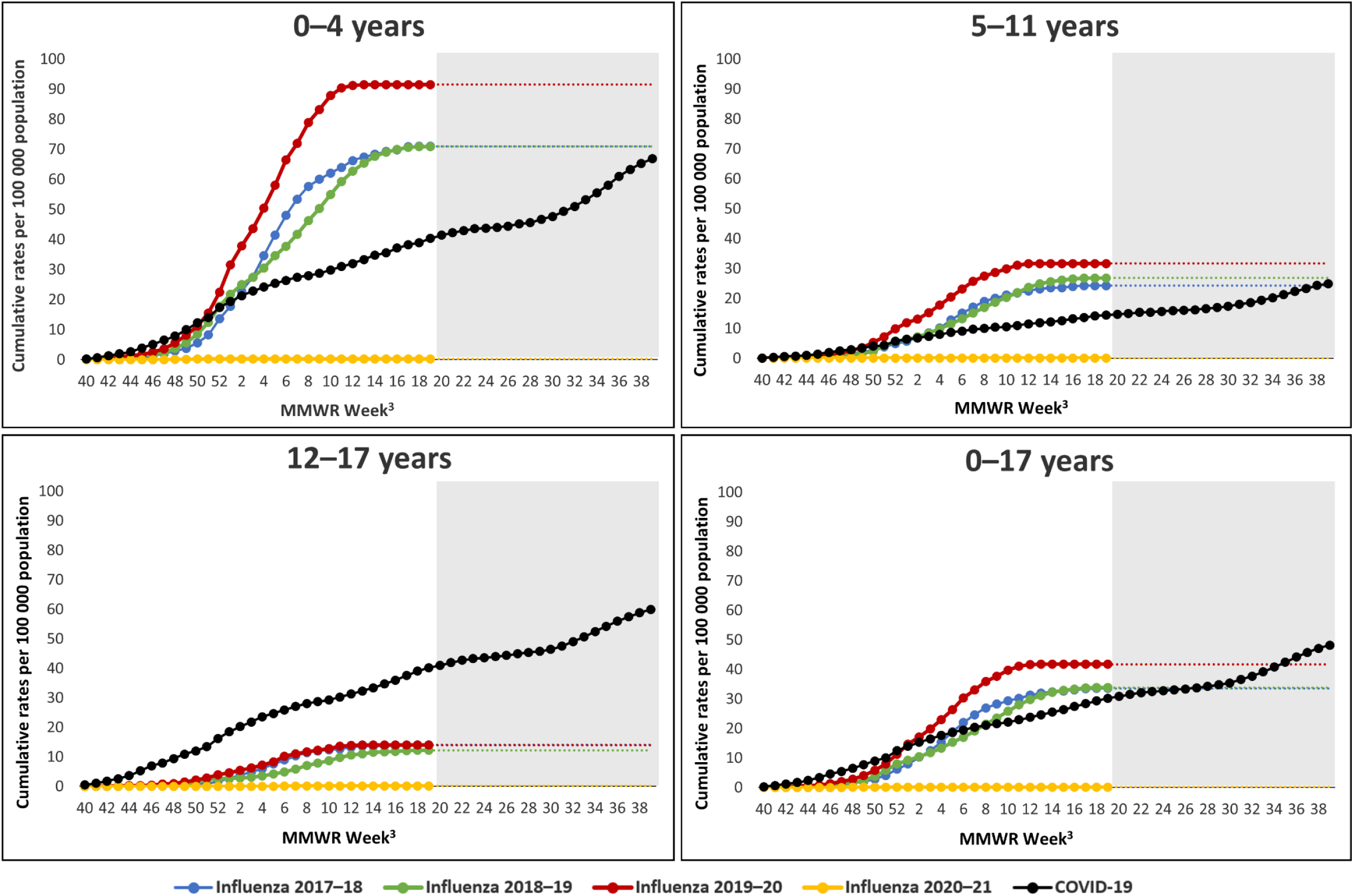
Cumulative influenza- and COVID-19-associated hospitalization rates per 100,000 children <18 years old, by age group – FluSurv-NET^1^ and COVID-NET^2^, 2017–2021. FluSurv-NET = Influenza Hospitalization Surveillance Network; COVID-NET = COVID-19-Associated Hospitalization Surveillance Network 1 Each season, FluSurv-NET surveillance is conducted from MMWR week 40 (around October 1) of one year to MMWR week 18 (around April 30) of the subsequent year. The grayed-out area on each panel indicates weeks during which FluSurv-NET surveillance was not conducted but COVID-NET surveillance was conducted. FluSurv-NET rate lines were extended beyond week 18 for ease of comparison with COVID-NET rate lines. 2 The COVID-NET surveillance period of October 2020–September 2021 begins at MMWR week 40 of year 2020 and ends at MMWR week 39 of year 2021. 3 MMWR Week 53 for year 2020 is combined with MMWR Week 52 for consistency with other years.

Weekly influenza-associated hospitalization rates peaked in February during all three seasons before the COVID-19 pandemic (peak weekly rate range: 2.4–4.0). Rates varied by influenza virus type across seasons (Figure 3). The highest weekly rate of COVID-19 (1.8) occurred in September 2021; additional peaks occurred in January and April 2021.

**Figure 3.**
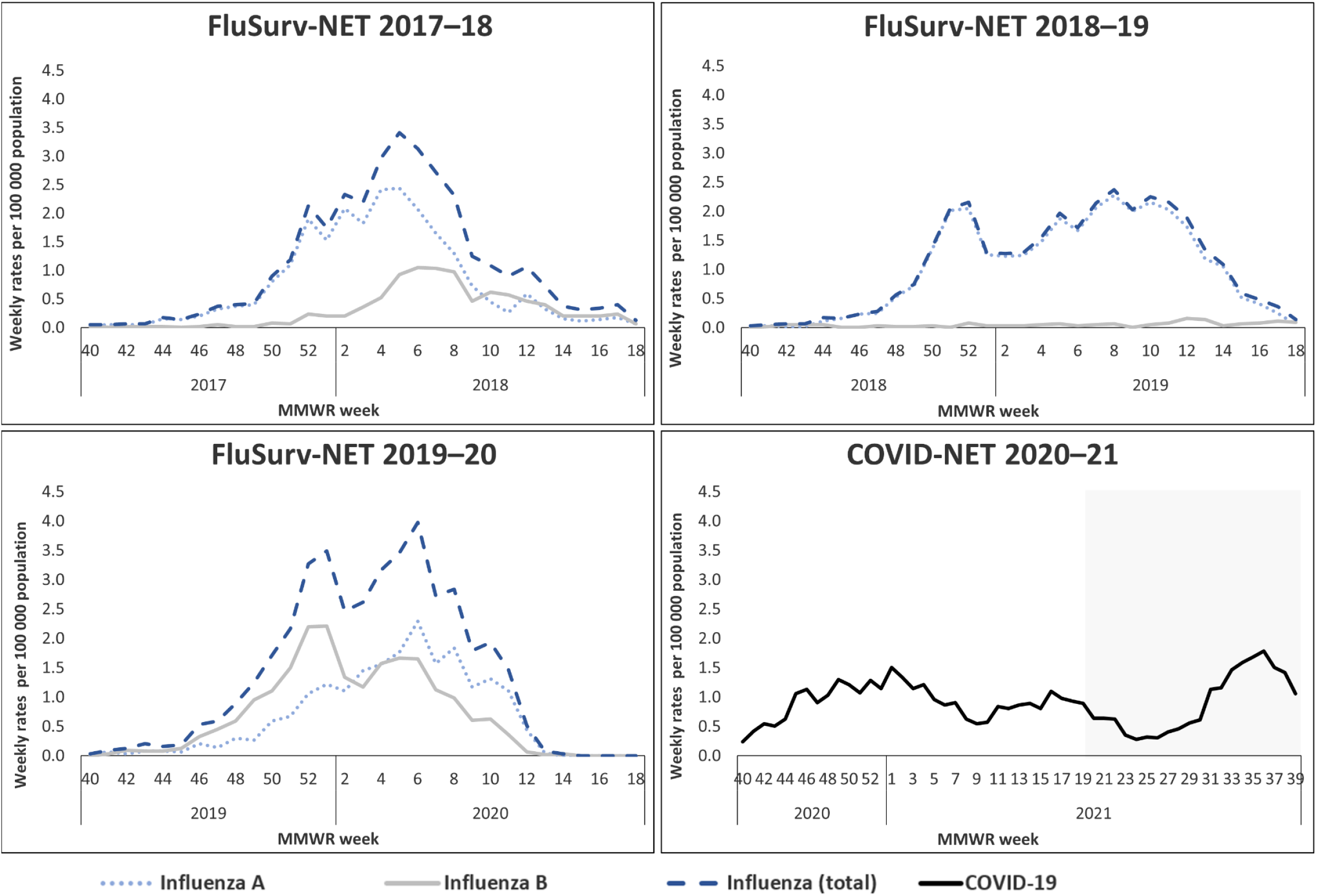
Weekly influenza- and COVID-19-associated hospitalization rates per 100 000 children <18 years old – FluSurv-NET^1^ and COVID-NET^2^. FluSurv-NET = Influenza Hospitalization Surveillance Network; COVID-NET = COVID-19-Associated Hospitalization Surveillance Network 1 FluSurv-NET data include the 2017–2018, 2018–2019, and 2019–2020 seasons. Each season, FluSurv-NET surveillance is conducted from MMWR week 40 (around October 1) of one year to MMWR week 18 (around April 30) of the subsequent year. 2 The COVID-NET surveillance period of October 2020–September 2021 begins at MMWR week 40 of year 2020 and ends at MMWR week 39 of year 2021. The grayed-out area on each figure indicates weeks during with FluSurv-NET surveillance was not conducted but COVID-NET surveillance was conducted.

Among 6774 children hospitalized with influenza during the 2017–18 through 2019–20 seasons and 3461 children hospitalized with COVID-19 during October 1, 2020–September 30, 2021, the median age was lower for influenza (3 years; interquartile range [IQR]: 1–7) than COVID-19 (9 years; IQR: 1–15) (Table 1). Other demographic characteristics were similar. Overall, 6564 children with influenza (96.9%) and 2760 children with COVID-19 (79.7%) had ≥ 1 symptom at admission. A higher proportion of children with influenza had ≥ 1 respiratory or febrile symptom compared to those with COVID-19 (95.6% vs 64.8%). Other common symptoms among children with influenza versus COVID-19 included nausea/vomiting (40.1% vs 34.4%), fatigue (29.1% vs 19.0%), and diarrhea (13.8% vs 15.8%) (Supplementary Table 1).

Overall, 3774 children hospitalized with influenza (55.7%) and 1857 children hospitalized with COVID-19 (53.7%) had ≥ 1 underlying medical condition (Table 2). Asthma/reactive airway disease, neurologic disorder, and obesity were the most prevalent underlying conditions for influenza and COVID-19. A higher proportion of children with influenza compared with COVID-19 had asthma (23.6% vs 16.3%) or chronic lung disease (6.0% vs 3.3%), but lower proportions had diabetes (1.2% vs 3.8%) or obesity (17.5% vs 35.0% among children ≥2 years old). The prevalence of obesity increased with age for COVID-19 and for influenza.

**Table 2.**
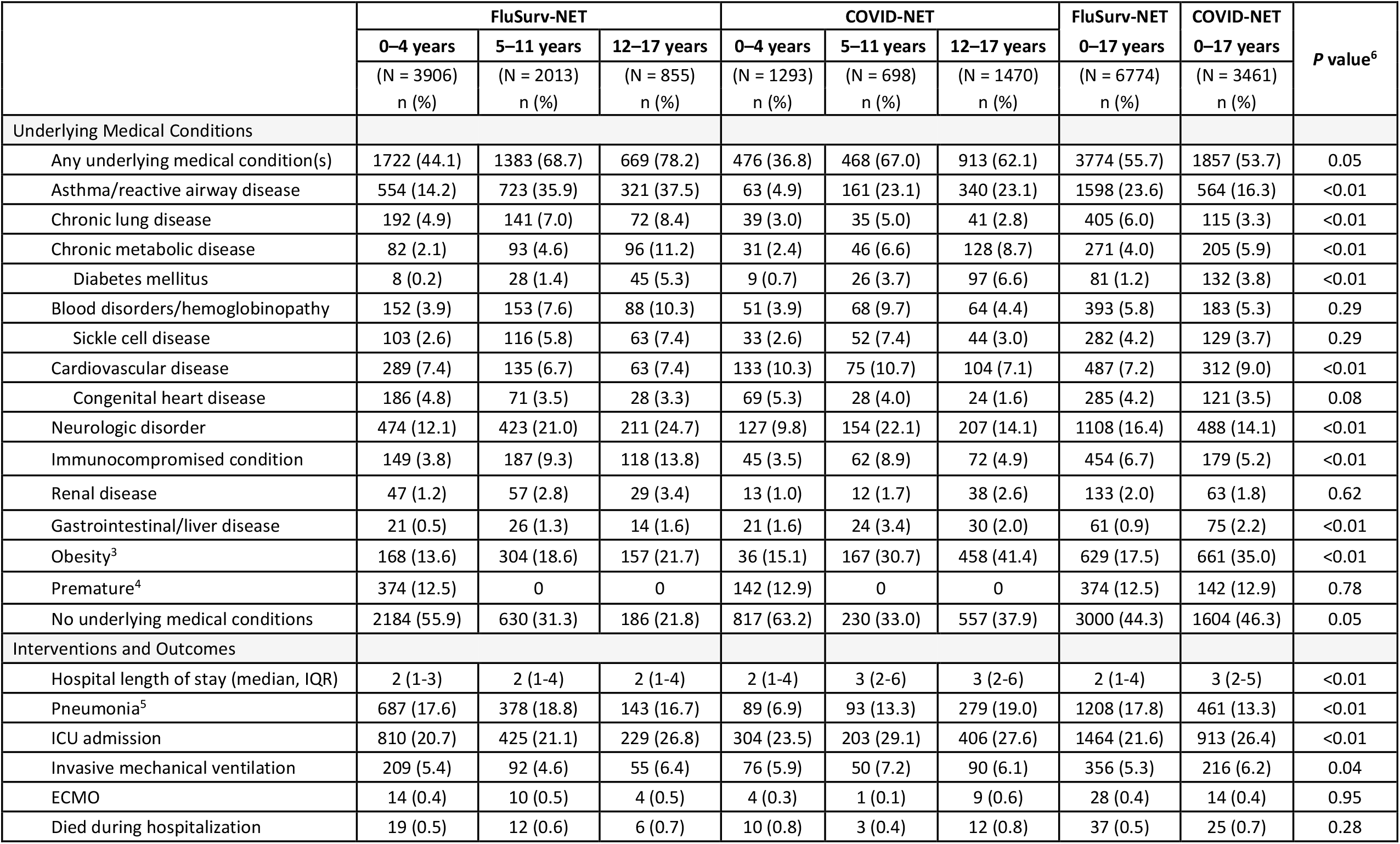
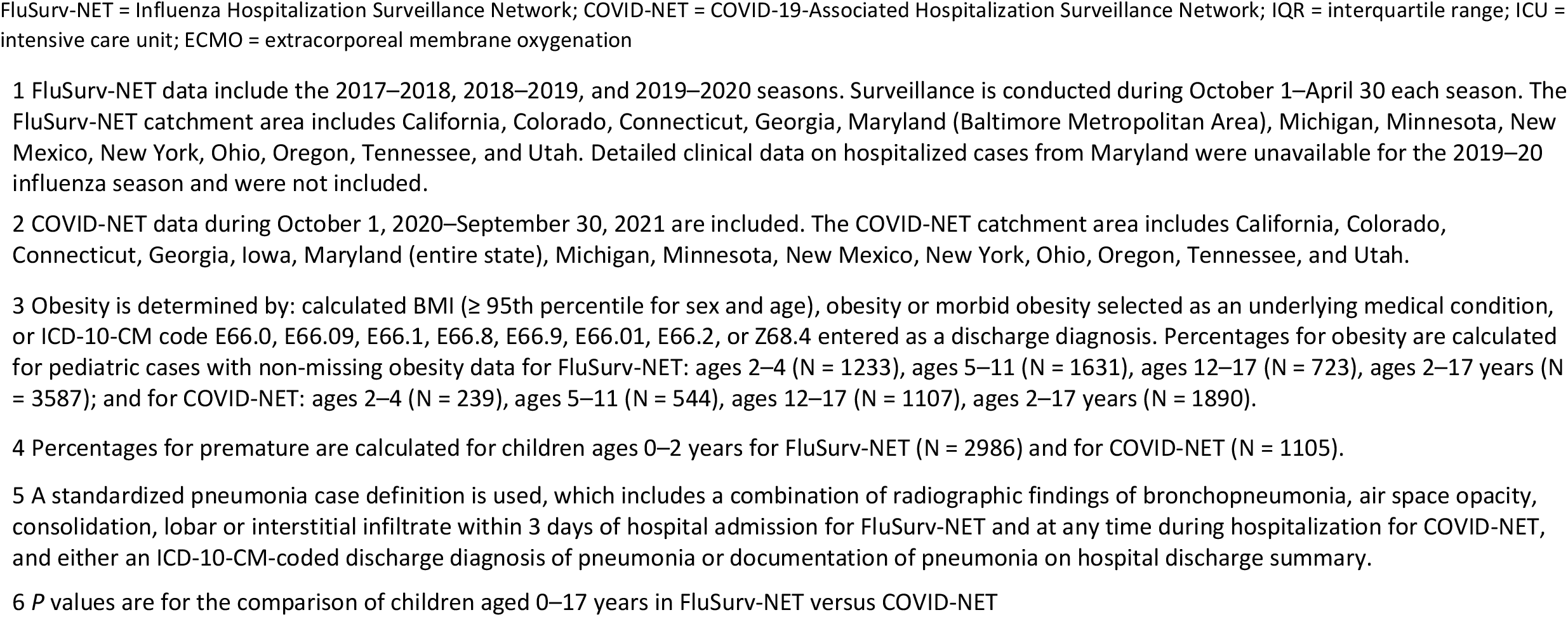
Underlying medical conditions, interventions, and outcomes of children <18 years old hospitalized with influenza or COVID-19– FluSurv-NET^1^ and COVID-NET^2^.

The median hospital length of stay was lower for children with influenza compared with COVID-19 (2 vs 3 days, p<0.01) (Table 2). A higher proportion of children with influenza compared with COVID-19 had pneumonia (17.8% vs 13.3%; p<0.01), but lower proportions required invasive mechanical ventilation (5.3% vs 6.2%; p=0.04) or ICU admission (21.6% vs 26.4%; p<0.01). The proportion of children with influenza vs COVID-19 who died during hospitalization was similar (0.5% vs 0.7%, p=0.28).

In sensitivity analyses, proportions experiencing severe outcomes were similar when examining all influenza or COVID-19-associated hospitalizations, influenza or COVID-19-associated hospitalizations among children with ≥ 1 symptom at admission (96.9% of 6774 influenza hospitalizations and 79.7% of 3461 COVID-19 hospitalizations), and COVID-19-associated hospitalizations with COVID-19 as the primary reason for admission (74.9% of COVID-19 hospitalizations) (Supplementary Table 2). Among COVID-19-associated hospitalizations, proportions with pneumonia or ICU admission increased modestly with increasing age when restricted to hospitalizations with ≥ 1 symptom at admission or COVID-19 as the primary reason for admission.

## Discussion

Using two large population-based surveillance platforms, we found that among children <18 years old, the COVID-19-associated hospitalization rate during one year of the ongoing COVID-19 pandemic (October 2020–September 2021) was higher than influenza-associated hospitalization rates observed during the three seasons prior to the pandemic, with differences observed by age group. Severe outcomes such as ICU admission, mechanical ventilation, and in-hospital death were generally similar among children with COVID-19 compared with influenza. Influenza has long been recognized as an important cause of severe respiratory illness in children in the United States and globally [1,18]. These data add to the growing literature demonstrating that COVID-19 is also an important cause of severe disease among children.

Prevention measures such as physical distancing, mask usage, and virtual learning likely contributed to historically low levels of influenza circulation and thus influenza-associated hospitalizations during the 2020–21 influenza season in the United States [19] and globally [20]. The COVID-19- and influenza-associated hospitalization rates among children during 2020–21 would likely have been higher without mitigation measures. While influenza activity typically displays a seasonal pattern, peaking during December–March in the northern hemisphere [15,19], COVID-19-associated hospitalizations occurred throughout 2020–2021 and no clear seasonality can be distinguished at this time. During the 2009 influenza A H1N1 pandemic (April 2009–April 2010), the H1N1pdm09 virus similarly circulated without distinct seasonality, resulting in spring and fall activity waves. Notably, rates of hospitalization and death associated with H1N1pdm09 were highest among children and younger adults compared with adults aged ≥65 years [21]. In subsequent seasons, H1N1pdm09 displayed a typical seasonal circulation pattern, with decreasing pediatric burden as population immunity levels increased. Given the relatively condensed annual period during which seasonal influenza viruses circulate, peak weekly hospitalization rates tended to be higher for influenza during 2017–2020 compared with COVID-19 during October 2020–September 2021, despite similar annual hospitalization rates. Co-circulation of influenza and SARS-CoV-2, along with other respiratory viruses, could exacerbate winter surges in pediatric hospitalizations, posing challenges to healthcare capacity [22,23].

While nearly all hospitalized children with influenza (95.6%) had a respiratory or febrile symptom, approximately one-third of children with COVID-19 did not. Large proportions of children with influenza or COVID-19 had non-respiratory symptoms including gastrointestinal symptoms and fatigue. Relying on respiratory or febrile symptoms alone to initiate influenza or SARS-CoV-2 testing could result in missed opportunities to detect infections. Respiratory virus testing can help distinguish between these viruses and guide treatment decisions and infection prevention practices [24–26]. There were also differences in the prevalence of underlying conditions among children with influenza versus COVID-19, which may in part be driven by differences in median age. The prevalence of obesity among children with COVID-19 was approximately double that of children with influenza. Notably, the proportion of children with COVID-19 who were obese (35%) was similar to findings from another study of hospitalized children with COVID-19 (32%) [27] and higher than the national obesity prevalence among children and adolescents 2–19 years old (22% in 2020) [28].

Our analysis and others demonstrate that both influenza and COVID-19 can cause severe disease in children [4,5]. Among hospitalized children, 22% with influenza and 26% with COVID-19 required ICU admission, and 5% with influenza and 6% with COVID-19 required mechanical ventilation. Another analysis of COVID-19 hospitalizations among children < 18 years old during July–August 2021, when the B.1.617.2 (Delta) variant of SARS-CoV-2 was predominant, showed similar findings with 29.5% requiring ICU admission and 7.9% requiring mechanical ventilation [27]. COVID-19 also can lead to multisystem inflammatory syndrome [29] and other longer-term sequelae [30]. Data on COVID-19 treatment in children are limited, but treatment may be indicated for hospitalized children who have an emergent or increasing need for supplemental oxygen [26]. To decrease the risk of severe complications, early influenza antiviral treatment is recommended for children with suspected influenza who are hospitalized or at higher risk for influenza-associated complications, including children <5 years old and those with underlying medical conditions [31].

Influenza vaccines are safe and effective at preventing hospitalizations among children, and were available to children ≥ 6 months old during all influenza seasons included in this analysis [6,32]. Based on national survey data, estimates of influenza vaccination coverage among children ≥ 6 months old ranged from 51–63% during the 2012–13 to 2018–19 seasons [33]. During the 2019–20 influenza season, influenza vaccination was estimated to avert 13,798 hospitalizations among children ≥ 6 months old [32]. Adolescents were the only children eligible to receive COVID-19 vaccination for a portion of the analytic period covered in this analysis: approvals differed over time by age (12–15 and 16–17 years) [7]. As of September 30, 2021, 47% of U.S. adolescents were considered up to date with all recommended COVID-19 doses [34]. COVID-19 vaccines are both safe and effective at preventing hospitalizations among adolescents and children 5–11 years old [35–38]. Increased COVID-19 vaccination coverage among children following the emergency use authorization for COVID-19 vaccines in children 5–11 years old in October 2021 may contribute to differences in the relative rates of pediatric COVID-19 vs influenza-associated hospitalizations over time.

Several limitations should be considered. First, children meeting COVID-NET and FluSurv-NET criteria may have been hospitalized for reasons other than COVID-19 [2] or influenza. This may have been more common for COVID-19 compared with influenza due to SARS-CoV-2 screening practices, which were universal among hospitalized patients at some facilities during certain time periods. While COVID-19 or influenza may not have been the primary reason for admission for all hospitalizations, such cases were included in rate calculations because use of a standard and consistent surveillance case definition allows for robust monitoring of trends over time. Among cases with influenza or SARS-CoV-2 incidentally identified, it is unclear what impact the infection ultimately had on the decision to hospitalize a patient, the hospitalization course, or in-hospital outcomes. In sensitivity analyses limited to hospitalizations where children had ≥ 1 symptom at admission or COVID-19 as the primary reason for admission, the proportions of children with severe outcomes were similar to proportions with severe outcomes when all hospitalizations were included. Second, COVID-19- and influenza-associated hospitalizations might have been missed because case identification was reliant on clinician-directed or facility-based testing practices and test availability, which varied across time and facilities. Under-detection of influenza was likely greater than for COVID-19 due to under-utilization of seasonal influenza testing [32]. Third, the impact of extensive mitigation measures during the COVID-19 pandemic, and differential availability of COVID-19 vaccines by age group and time period could not be measured, and likely affects the comparison of influenza versus COVID-19-associated hospitalization rates. Fourth, only deaths occurring during hospitalizations were captured, which may miss deaths associated with COVID-19 or influenza occurring after hospital discharge [39]. Fifth, the COVID-NET and FluSurv-NET catchment areas include approximately 9–10% of the U.S. population and findings may not be generalizable to the entire country. Last, this analysis assessed COVID-19-associated hospitalization rates during a single year of the ongoing COVID-19 pandemic and did not capture all rate fluctuations that have occurred due to the changing epidemiology of SARS-CoV-2, including the emergence of variants of concern. The omicron variant of SARS-CoV-2 emerged rapidly during December 2021 and resulted in a peak weekly hospitalization rate approximately five times as high as the peak hospitalization rate during the period of Delta variant predominance among children 0–4 years old, a group not yet eligible for COVID-19 vaccination [40].

Influenza and SARS-CoV-2 are important causes of hospitalization and severe outcomes among children. Vaccines can help prevent illness and attenuate disease severity for both influenza and COVID-19. Prevention and mitigation measures, including vaccination of all eligible persons, are crucial to protect children, including those who are not yet eligible for COVID-19 vaccination or too young for influenza vaccination. Without such measures, co-circulation of influenza virus and SARS-CoV-2, along with other respiratory viruses, could exacerbate surges in hospitalizations and overwhelm healthcare capacity, particularly during the winter months.

## Data Availability

All data produced in the present study are available upon reasonable request to the authors

## Acknowledgements

Gretchen Rothrock, Jeremy Roland, Joelle Nadle, Pam Kirley (California Emerging Infections Program); Isaac Armistead, Sarah McLafferty (Colorado Department of Public Health and Environment); Adam Misiorski, Amber Maslar, Carol Lyons, Daewi Kim, Maria Correa, Paula Clogher, Tessa Carter (Connecticut Emerging Infections Program, Yale School of Public Health); Allison Roebling, Annabel Patterson, Asmit Joseph, Chandler Surell, Emily Fawcett, Grayson Kallas, Jana Manning, Jeremiah Williams, Katelyn Ward, Marina Bruck, Rayna Ceaser, Siyeh Gretzinger, Stephanie Lehman, Taylor Eisenstein (Georgia Emerging Infections Program, Georgia Department of Public Health, Atlanta Veterans Affairs Medical Center); Suzanne Segler (Division of Infectious Diseases, Emory University School of Medicine, Georgia Emerging Infections Program, Georgia Department of Public Health); Alicia Brooks, Cindy Zerrlaut, David Blythe, Elisabeth Vaeth (Maryland Department of Health); Michelle Wilson, Rachel Park (Maryland Emerging Infections Program, The Johns Hopkins Bloomberg School of Public Health); Alexander Kohrman, Chloe Brown, Jim Collins, Justin Henderson, Shannon Johnson, Sierra Peguies-Khan, Sue Kim, Val Tellez Nunez (Michigan Department of Health and Human Services); Alexandra Warzecha, Alison Babb, Amanda Gordon, Austin Bell, Claire Henrichsen, Cynthia Kenyon, Elizabeth Corey, Emily Holodnick, Emma Contestabile, Erica Bye, Grace Hernandez, Jackie Johnson, Jade Van Kley, Jennifer Gilbertson, Jill Reaney, Kathryn Como-Sabetti, Kayla Bilski, Kelli Aarstad, Kieu My Phipps, Lisa Nguyen, Natalie Bullis, Richie Xu, Samantha Siebman (Minnesota Department of Health); Chelsea McMullen, Emily Hancock, Jasmyn Sanchez, Kathy Angeles, Mayvilynne Poblete, Melissa Christian, Melissa Judson, Nancy Eisenberg, Sarah Khanlian, Sarah Lathrop, Sarah Shrum Davis, Sunshine Martinez, Wickliffe Omondi, Yadira Salazar-Sanchez, Yassir Talha (New Mexico Emerging Infections Program); Murtada Khalifa (CDC Foundation, New Mexico Department of Health); Cory Cline, Daniel Sosin (New Mexico Department of Health); Adam Rowe, Grant Barney, Kerianne Engesser, Suzanne McGuire (New York State Department of Health); Christina Felsen, Christine Long, Maria Gaitán, RaeAnne Kurtz, Sophrena Bushey, Thomas Peer, Virginia Cafferky (University of Rochester School of Medicine and Dentistry); Ann Salvator, Julie Freshwater (Ohio Department of Health); Ama Owusu-Dommey, Nasreen Abdullah, Nicole West, Sam Hawkins (Public Health Division, Oregon Health Authority); Anise Elie, Bentley Akoko, Danielle Ndi, John Ujwok, Karen Leib, Kathy Billings, Katie Dyer, Manideepthi Pemmaraju, Terri McMinn, Tiffanie Markus, Victoria Umutoni (Vanderbilt University Medical Center); Amanda Carter, Andrea George, Andrea Price, Andrew Haraghey, Ashley Swain, Caitlin Shaw, Ian Buchta, Jake Ortega, Laine McCullough, Mary Hill, Ryan Chatelian, Tyler Riedesel (Salt Lake County Health Department); Sonja Nti-Berko, Alvin Shultz (Emerging Infections Program); Mimi Puckett (Council of State and Territorial Epidemiologists).

## Disclaimer

The conclusions, findings, and opinions expressed by the authors do not necessarily reflect the official position of the United States Department of Health and Human Services, the United States Public Health Service, the Centers for Disease Control and Prevention, or the authors’ affiliated institutions.

## Funding

This work was supported by the Centers of Disease Control and Prevention through an Emerging Infections Program cooperative agreement (grant CK17–1701) and through a Council of State and Territorial Epidemiologists cooperative agreement (grant NU38OT000297–02–00).

## Potential conflicts of interest

Dr. Evan Anderson reports grants from Pfizer, grants from Merck, grants from PaxVax, grants from Micron, grants from Sanofi-Pasteur, grants from Janssen, grants from MedImmune, grants from GlaxoSmithKline, personal fees from Sanofi-Pasteur, personal fees from Pfizer, personal fees from Medscape, personal fees from Kentucky Bioprocessing, Inc, personal fees from Sanofi-Pasteur, outside the submitted work. Dr. William Schaffner reports personal fees from VBI Vaccines, outside the submitted work.

---

**Supplementary Table 1.** Other symptoms at admission among children <18 years old hospitalized with influenza or COVID-19– FluSurv-NET^1^ and COVID-NET^2^.

|  | FluSurv-NET 2017-2018 |  |  | COVID-NET |  |  | FluSurv-NET | COVID-NET |
| --- | --- | --- | --- | --- | --- | --- | --- | --- |
|  | 0–4 years | 5–11 years | 12–17 years | 0–4 years | 5–11 years | 12–17 years | 0–17 years | 0–17 years |
|  | (N = 1251)<br>n (%) | (N = 613)<br>n (%) | (N = 308)<br>n (%) | (N = 1293)<br>n (%) | (N = 698)<br>n (%) | (N = 1470)<br>n (%) | (N = 2172)<br>n (%) | (N = 3461)<br>n (%) |
| Other symptom(s)<sup>3</sup> |  |  |  |  |  |  |  |  |
| Abdominal pain<sup>4</sup> | N/A | N/A | N/A | 44 (23.4) | 228 (32.7) | 354 (24.1) | N/A | 626 (26.6) |
| Chest pain<sup>4</sup> | 9 (3.3) | 46 (7.5) | 68 (22.1) | 5 (2.7) | 61 (8.7) | 256 (17.4) | 123 (10.3) | 322 (13.7) |
| Conjunctivitis | 29 (2.3) | 8 (1.3) | 2 (0.6) | 40 (3.1) | 33 (4.7) | 14 (1.0) | 39 (1.8) | 87 (2.5) |
| Diarrhea | 203 (16.2) | 57 (9.3) | 39 (12.7) | 224 (17.3) | 115 (16.5) | 209 (14.2) | 299 (13.8) | 548 (15.8) |
| Fatigue | 329 (26.3) | 199 (32.5) | 104 (33.8) | 180 (13.9) | 172 (24.6) | 305 (20.7) | 632 (29.1) | 657 (19.0) |
| Headache<sup>4</sup> | 28 (10.2) | 104 (17.0) | 85 (27.6) | 18 (9.6) | 135 (19.3) | 332 (22.6) | 217 (18.2) | 485 (20.6) |
| Hemoptysis | N/A | N/A | N/A | 1 (0.1) | 2 (0.3) | 16 (1.1) | N/A | 19 (0.5) |
| Loss of smell<sup>4</sup> | N/A | N/A | N/A | 0 | 8 (1.1) | 67 (4.6) | N/A | 75 (3.2) |
| Loss of taste<sup>4</sup> | N/A | N/A | N/A | 2 (1.1) | 12 (1.7) | 72 (4.9) | N/A | 86 (3.7) |
| Myalgia<sup>4</sup> | 33 (12.0) | 102 (16.6) | 81 (26.3) | 10 (5.3) | 68 (9.7) | 203 (13.8) | 216 (18.1) | 281 (11.9) |
| Nausea/vomiting | 471 (37.6) | 273 (44.5) | 127 (41.2) | 384 (29.7) | 314 (45.0) | 491 (33.4) | 871 (40.1) | 1189 (34.4) |
| Rash | 70 (5.6) | 25 (4.1) | 12 (3.9) | 124 (9.6) | 58 (8.3) | 35 (2.4) | 107 (4.9) | 217 (6.3) |

FluSurv-NET = Influenza Hospitalization Surveillance Network; COVID-NET = COVID-19-Associated Hospitalization Surveillance Network
1 FluSurv-NET data include hospitalizations during October 1, 2017–April 30, 2018.
2 COVID-NET data include hospitalizations during October 1, 2020–September 30, 2021.
3 Other symptoms collected by COVID-NET were: myalgia, chest pain, loss of taste, loss of smell, diarrhea, conjunctivitis, fatigue, headache, rash, nausea/vomiting, abdominal pain, and hemoptysis; for children <2 years old, symptoms also included apnea, cyanosis, decreased vocalization/stridor, dehydration, hypothermia, inability to eat/poor feeding, and lethargy. Other symptoms collected by FluSurv-NET in the 2017–18 season were: myalgia, chest pain, diarrhea, conjunctivitis, fatigue, headache, nausea/vomiting, and rash. Denominators for percentages for other symptoms for FluSurv-NET are for the 2017–2018 season for ages 0–4 (N = 1251), ages 5–11 (N = 613), ages 12–17 (N = 308), and ages 0–17 years (N = 2172). No additional symptoms were collected for FluSurv-NET in the 2018–19 or 2019–20 seasons.
4 Symptom data on abdominal pain, chest pain, headache, loss of smell, loss of taste, and myalgia were collected for children of all ages but reported only among children aged ≥ 3 years (for FluSurv-NET: N = 274 children ages 3–4 and N = 1195 ages 3–17 years; for COVID-NET: N = 188 ages 3–4 and N = 2356 ages 3–17 years).

**Supplementary Table 2.**
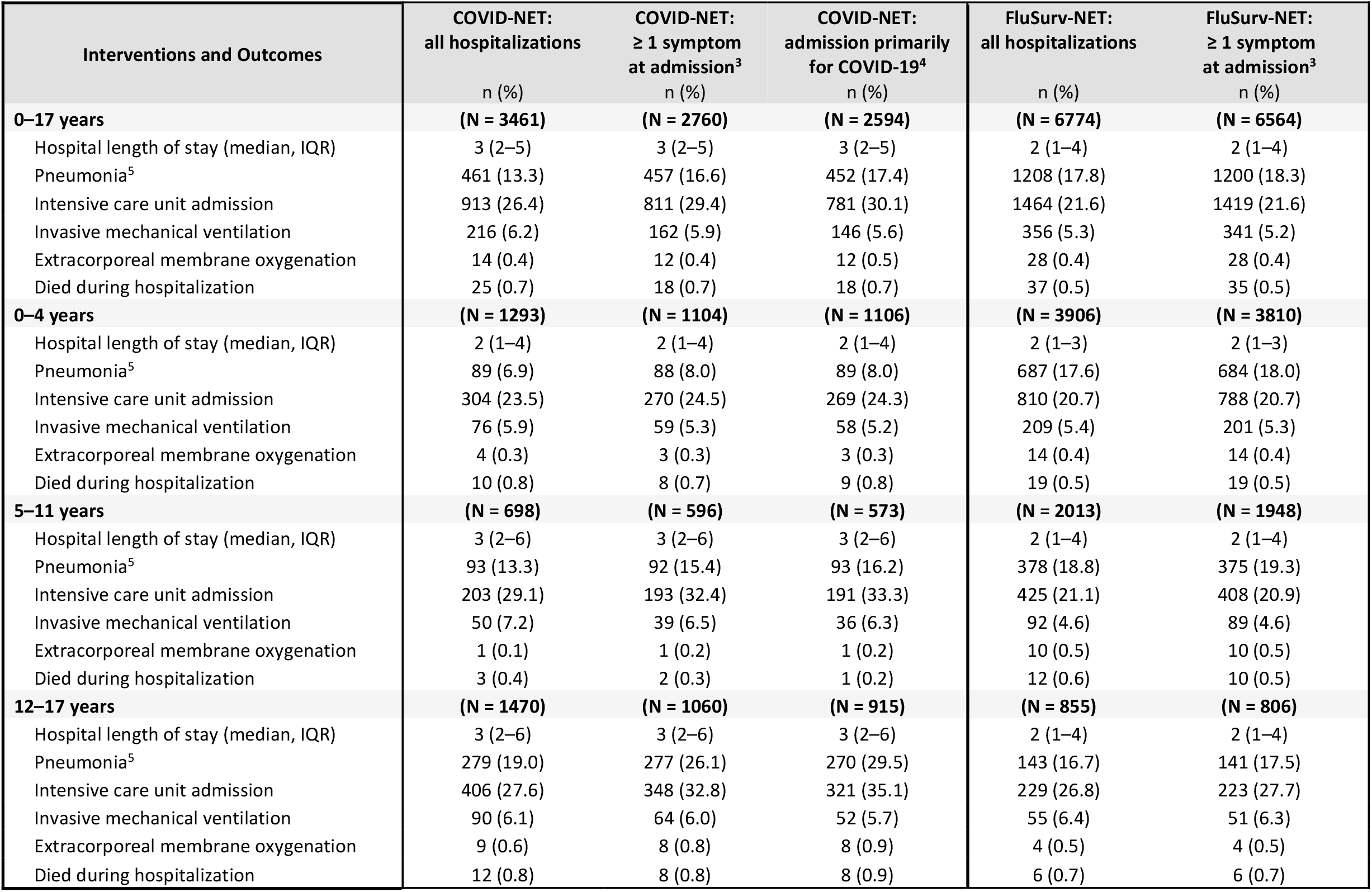
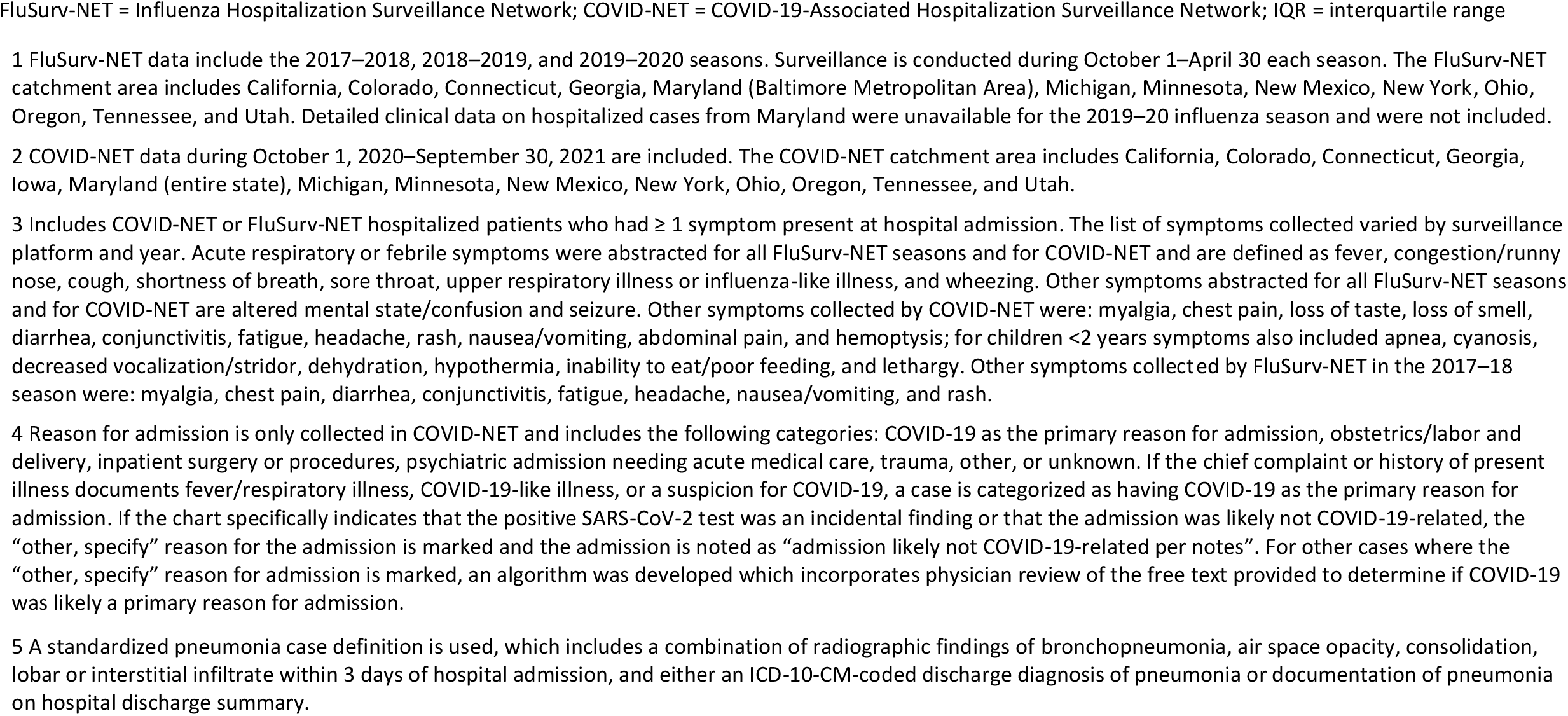
Interventions and outcomes among children <18 years old hospitalized with influenza or COVID-19 by symptom status and reason for admission, FluSurv-NET^1^ and COVID-NET^2^.

**Supplementary Table 3.** Annual cumulative influenza- and COVID-19-associated hospitalization rates per 100,000 children <18 years old, by age group and season – FluSurv-NET^1^ and COVID-NET^2^.

| Surveillance System/Season | Age Group (years) | Cumulative Hospitalization Rate (95% CI) |
| --- | --- | --- |
| FluSurv-NET 2017–18 | 0–4 | 70.96 (67.02–74.89) |
| FluSurv-NET 2018–19 | 0–4 | 70.91 (66.97–74.86) |
| FluSurv-NET 2019–20 | 0–4 | 91.50 (86.99–96.01) |
| FluSurv-NET 2020–21 | 0–4 | 0.29 (0.04–0.55) |
| COVID-NET Oct 2020–Sep 2021 | 0–4 | 66.81 (63.17–70.45) |
| FluSurv-NET 2017–18 | 5–11 | 24.29 (22.37–26.21) |
| FluSurv-NET 2018–19 | 5–11 | 26.81 (24.79–28.84) |
| FluSurv-NET 2019–20 | 5–11 | 31.66 (29.46–33.87) |
| FluSurv-NET 2020–21 | 5–11 | 0.00 |
| COVID-NET Oct 2020–Sep 2021 | 5–11 | 24.96 (23.11–26.81) |
| FluSurv-NET 2017–18 | 12–17 | 14.05 (12.48–15.61) |
| FluSurv-NET 2018–19 | 12–17 | 12.19 (10.72–13.65) |
| FluSurv-NET 2019–20 | 12–17 | 13.95 (12.39–15.52) |
| FluSurv-NET 2020–21 | 12–17 | 0.14 (0.00–0.29) |
| COVID-NET Oct 2020–Sep 2021 | 12–17 | 59.94 (56.88–63.00) |
| FluSurv-NET 2017–18 | 0–17 | 33.52 (32.11–34.93) |
| FluSurv-NET 2018–19 | 0–17 | 33.80 (32.38–35.22) |
| FluSurv-NET 2019–20 | 0–17 | 41.73 (40.15–43.31) |
| FluSurv-NET 2020–21 | 0–17 | 0.12 (0.04–0.21) |
| COVID-NET Oct 2020–Sep 2021 | 0–17 | 48.18 (46.57–49.78) |

FluSurv-NET= Influenza Hospitalization Surveillance Network; COVID-NET= COVID-19-Associated Hospitalization Surveillance Network
1 Each season, FluSurv-NET surveillance conducts surveillance from MMWR week 40 (around October 1) of one year to MMWR week 18 (around April 30) of the subsequent year.
2 The COVID-NET surveillance period of October 2020–September 2021 includes MMWR week 40 of year 2020 to MMWR week 39 of year 2021.

## Notes

### Author Declarations

FluSurv-NET and COVID-NET surveillance activities were reviewed by CDC and were conducted consistent with applicable federal law and CDC policy (e.g., 45 CFR. Part 46.102(l)(2), 21 CFR part 56; 42 USC. Section 241(d); 5 USC Section 552a; 44 USC Section 3501 et seq). Sites participating in FluSurv-NET and COVID-NET obtained human subjects and ethics approvals from their respective state and local health department and academic partner Institutional Review Boards as needed.

